# Multiple Long-Term Conditions (MLTC) and the environment: A scoping review

**DOI:** 10.1101/2022.06.28.22276984

**Authors:** Hajira Dambha-Miller, Sukhmani Cheema, Nile Saunders, Glenn Simpson

**Author notes:** **Correspondence:** Dr Hajira Dambha-Miller, Primary Care Research Centre, The University of Southampton, Aldermoor Health Centre, Aldermoor Close, United Kingdom, SO16 5ST.

## Abstract

**Background:** Multiple Long Term conditions (MLTC), the coexistence of two or more health conditions, is a major health care challenge associated with high service utilisation and expenditure. Once established, the trajectory to an increased number and severity of conditions, hospital admission, increased social care need and mortality is multifactorial. The role of wider environmental determinants in the MLTC sequelae is unclear.

**Aim:** The aim of this review was to summarise and collate existing evidence on environmental determinants on established MLTC.

**Methods:** A comprehensive search of Medline, Embase, Cochrane, CINAHL and Bielefeld Academic Search Engine (BASE) from inception to 4th June 2022 in addition to grey literature. Two authors independently screened and extracted papers. Disagreements were resolved with a third author.

**Results:** The search yielded 9,079 articles of which 12 were included. Five studies considered the effect of urban built environments and neighbourhood characteristics on MLTC. Two studies examined both the built and social environments. The social environment was considered by four studies. One study examined the natural environment. Evidence of correlations between some environmental determinants and increased or decreased risks of MLTC were found, including the quality of internal housing/living environments, exposure to airborne environmental hazards and a beneficial association with socially cohesive, accessible and greener neighbourhood environments.

**Conclusions:** Only 12 relevant papers were identified, with the majority focused on the built and social environments. Overall, the review uncovered very limited evidence and this finding indicates a need for further research to understand the role of environmental determinants in MLTC.

## INTRODUCTION

Multiple Long Term conditions (MLTC) are a rapidly growing health care challenge due to increased longevity and a rising prevalence of chronic health conditions. Where two or more health conditions coexist in this way, medical and social care needs are greater, service usage and expenditure are also higher (1,2). The prevalence of MLTC will grow significantly over the next decade, with the number of people aged 65-plus with two or more diseases increasing from 54% in 2015 to a projected 67% by 2035 (3). It is estimated that by 2035 around 17% of the UK’s population will have four or more chronic conditions, which is almost double the current prevalence (9·8%) (4). There are also substantial health and social costs for individuals with MLTC, who are likely to experience more severe illness and complication rates, increased physical and mental disability, lower quality of life and risk of social vulnerability including homelessness, unemployment and poverty (5–7).

A growing body of evidence has highlighted the multifactorial causes of MLTC including wider health, social and environmental determinants. Environmental determinants are those that occur in natural, built or social environments (8).The built environment refers to ‘the characteristics (objective and subjective) of a physical environment in which people live, work and play, including schools, workplaces, homes, communities, parks/recreation areas, green (i.e., visible grass, trees and other vegetation) and blue spaces (i.e., visible water)’ (9). The quality of the natural and built environment can have either a deleterious or beneficial impact on the health of an individual or the wider population health of a community (10). Much of the literature focuses on the harmful effects or health risks associated with poor quality natural and built environments (11). Understood in this context, environmental determinants of health are those attributed to ‘exposure to pollution and chemicals (e.g., air, water, soil, products), physical exposures (e.g., noise, radiation), the built environment (e.g., housing, land-use, infrastructure), other anthropogenic changes (e.g., climate change, vector breeding places), related behaviours and the work environment’ (12). As Pruss-Ustin (2017) (12) comment, these human-made or modified determinants of health that are external to the person, can be ameliorated or prevented by policy interventions ‘either with almost immediate effect, or with longer term transformations’. Interventions to address environmental determinants in relation to the built and physical environment typically focus on a range of policy areas, which directly or indirectly affect health and wellbeing of individuals and populations (13). ‘Direct’ environmental determinants are those ‘traditionally associated with infrastructure planning and environmental health’ (e.g., air quality, noise pollution and traffic-related factors) and where health impacts are ‘quantifiable and causal effects can be attributed’ (14). Indirect determinants, which can be more difficult to measure are ‘the ways in which built environment features and their design can influence the feelings and behaviour of individuals and populations. For example, perceptions of the local area, social connections, accessibility and physical activity levels’ (14). These forms of indirect determinants are associated with social determinants of health (15). Importantly, these built and physical environment determinants of health are often spatially concentrated (i.e., spatial inequalities in health), with deprived areas, populations and communities being more disproportionately affected as a result of living in degraded and/or poorly designed physical spaces and environments (16).

In MLTC, there is limited evidence on the role of environmental determinants, especially in relation to the trajectories of those with established MLTC towards declining health, mortality and increased care needs. The limited studies that have been published in this field have tended to focus on biological or clinical determinants such as weight changes or variations in blood markers (12,15). This is owing to the fact that these determinants are more easily available and relatively straightforward to measure, compared to environmental determinants.

Understanding an individual’s MLTC trajectory and subsequent outcomes in relation to their immediate environments could provide opportunities to identify key events in their life course at different geographical scales. In turn, this could help identify populations at risk of worsening MLTC trajectories by localisation (e.g. within a neighbourhood context) and exposures within their surrounding environment, alongside health and social determinants. This is essential in designing effective solutions and influencing response mechanisms within health services as it can indicate locations in need of interventions and allocation of available resources. Accordingly, in this review we collate and summarise evidence on environmental determinants that have been examined in relation to established MLTC.

## METHODS

### Review approach

The study followed the Preferred Reporting Items for Systematic Reviews and Meta-Analyses (PRISMA) guidelines for scoping reviews (17). The scoping review method allows for a rapid mapping of key existing research and emerging evidence in this field (18).

### Search strategy

Systematic electronic searches were conducted from database inception to 4th June 2022 on Medline, EMBASE, The Cochrane Library, CINAHL and Bielefeld Academic Search Engine (BASE). For searches of electronic databases, free-text and MeSH terms were used in relation to ‘multimorbidity’ and ‘environmental’. Detailed search terms are shown in the Appendix (Table 1). Manual searching of bibliographies were also conducted. The views of topic experts were also sought to identify additional sources.

### Inclusion/exclusion criteria

Articles were eligible for inclusion if published in the English language and related to adults aged over 18 years with MLTC and included mention of any environmental determinants.

Quality assessment criteria are not a priority for scoping reviews (18), therefore extracted articles were not excluded on this basis.

### Study selection and data extraction

Titles and abstracts were screened, with each article assessed for relevance according to the inclusion criteria. Full-text articles were retrieved. Both screening and data extraction were conducted independently by two reviewers and disagreement resolved by discussion. A data charting form was used, to collate the studies and identify key characteristics (Appendix, Table 2). Reviewers extracted the article reference and date, the stated aim of the study, methods, results, and variables or indicators of ‘environmental factors,’ key findings. Any disagreement between reviewers was resolved with a third reviewer.

### Summarising and analysis

The data gathered by the review were iteratively synthesised descriptively, using counts to summarise article characteristics (that set out the number, type, and quality of studies extracted), the data charting technique and interpretation of the findings by sifting and sorting material (19). The main environmental factors identified were collated and presented below.

## RESULTS

### Screening process

In total 9,079 articles were identified. Following title and abstract screening and removal of duplicates using the Rayyan review tool, 138 articles underwent full-text screening, which resulted in a further 126 articles excluded from the review. Reasons for exclusion were:

- the article did not analyse environmental determinants;
- the article did not specifically focus on the topic of multiple long term conditions;
- the article was not related to the study population of adults aged18+.

A final total of 12 articles were included in the review. A flowchart illustrating the screening process, including the reasons for exclusion, is shown in the supplementary material (Figure 1).

**Figure 1-.**
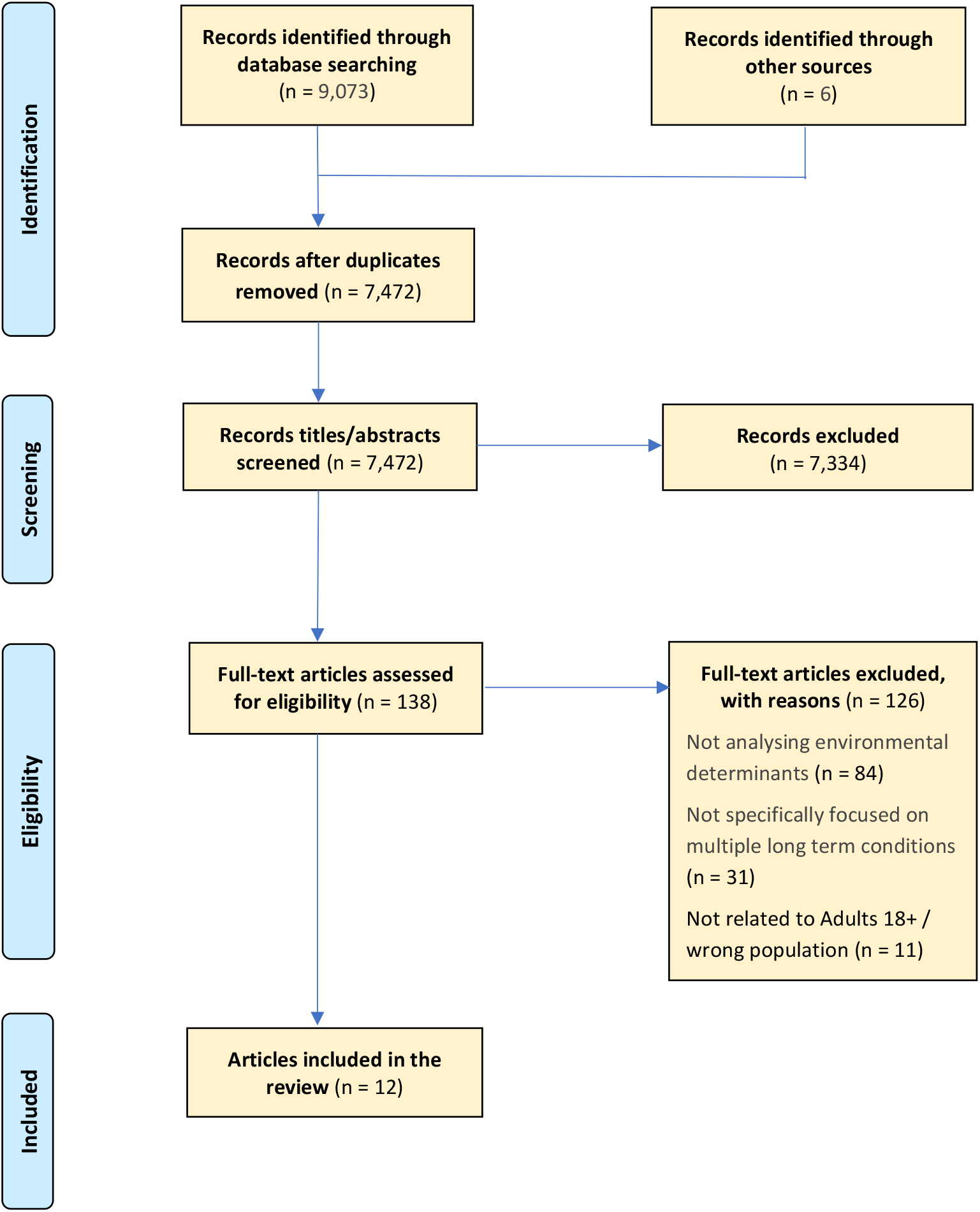
Adapted PRISMA Flow Chart Explaining the study’s documentary screening and inclusion/exclusion process.

### Characteristics of included studies

Studies were located in a variety of countries and international regions, most frequently the USA (n=3), followed by Australia, Canada (n=2) and UK, South Africa, Sub-Saharan Africa, North America-Europe-Australasia and no specified geography (n=1). The main study settings identified were community/neighbourhood (n=6), community and people at home (n=3), no specific setting (n=2) and low-income housing developments (n=1). The main age category found in the studies was Aged 18 and over (n=7), with the remaining studies focused on middle and older aged populations of 45 years and above, while one study did not specify a specific age range. Wide variation in the sample size was found among the studies, ranging from a sample of 20 (in a mixed study) to a study population of 408,111 (in a quantitative research). The main study type was quantitative (n=6), followed by both mixed methods (n=3) and reviews (n=3).

The characteristics of included articles are summarised in Table 2.

### Summary of environmental factors

Studies discussed a range of environmental determinants and possible associations with MLTC, which included those relating to the natural, built and/or social environments.

#### Urban built environment

Five studies considered various aspects relating to the effect of urban environments and neighbourhood characteristics on multimorbidity. One study (I) examined environmental factors which supported the ‘maintenance of functional ability’. This study highlighted the importance of physical proximity in urban contexts for the cohort aged 85 and over with MLTC, concluding that ‘proximity to local grocery shops may support their functional ability’ to undertake grocery shopping ‘over a five-year period’.

Another study (II) considered whether the built and urban environments, including its condition, configuration and layout inhibited or facilitated walking (‘walkability’), along with accessibility to recreation parks in urban neighbourhoods. Findings indicated that ‘living in a walkable neighbourhood and having higher park accessibility is associated with lower odds of hypertension, especially for lower income individuals.’ The authors suggested ‘an integrated population health approach that considers multimorbidity as a result of exposure to car-dependent areas and the lack of green spaces.’

A study examined (III) exposure to outdoor air pollution and/or hazards relating to air quality and associations with the development of chronic conditions in low to medium income countries (LMICs). Results suggested that compared with developed states, populations in LMICs are potentially more exposed to ‘poorer air quality and increased risk of multimorbidity’ because of factors including ‘overpopulation and rapid urbanisation coupled with developing industrialisation’, as well as higher usage of and reliance on ‘wood fuel’ due to higher levels of poverty.

One study (IV) examined conditions within household environments, in particular the effect of ‘housing-related environmental exposures’ on health in low-income households. Using ‘binary indexes and a summed index’ covering six household exposures (these were: mold, combustion by-products, second-hand smoke, chemicals, pests, and inadequate ventilation), the study identified ‘significant clustering of effects in [the] housing site for 4 of the 6 indexes: pests, combustion by-products, mold, and ventilation.’ A major weakness reported was that the ‘study design was cross-sectional and thus could not determine the causal relationship between exposures and health,’ including any association with MLTC.

Another study (V) sought to determine ‘evidence gaps to building capacity in supportive housing health care research about chronic diseases and multimorbidity, identify ongoing strains and offer potential solutions’. It presented a ‘multifaceted framework’ of eight priorities for action, including ‘scaling up environmental determinants to cover residential spaces’; implementing acute methodologies for indoor environment risk assessment;’ and ‘increasing clinical research applicability into built environment studies’. The authors recommended that future policy interventions should explore ‘approaches to strengthen health and care delivery through housing design solutions for people living with chronic morbidity.’

#### The built and social environments

Two studies examined both the built and social environments. One study (VI) explored the ‘association between neighborhood characteristics [defined as five ‘contextual neighborhood factors’ of: neighborhood-level crime, accessibility to health care services, availability of green spaces, neighborhood obesity, and fast-food availability] and type 2 diabetes (T2D) comorbidity in serious mental illness (SMI). Results showed that individuals with SMI living in high crime areas ‘had 2.5 times increased odds of reporting T2D comorbidity’ compared with those in ‘lower crime rate areas’. No causal association was found in relation to the other four neighbourhood characteristics variables.

Another study (VII) explored self-rated health and its association with multiple types of perceived environmental hazards, including participant’s perceptions of neighbourhood ‘environmental hazards (e.g., air quality, odors and noise)’, self-reported ‘aspects of the social environment (e.g., feeling safe, neighborhood crime, social cohesion)’ as well as ‘culture-related stressors (e.g., immigration status, language stress, ethnic identity)’. Findings highlighted ‘negative perceptions of environmental hazards and reported cultural stressors were significantly associated with fair/poor self-rated health among residents in a low-income majority-minority community’. Social cohesion was found to have a ‘beneficial association with self-rated health’.

#### The social environment

The social environment was considered by four studies. Housing and wider living conditions were the focus of one study (VIII), which examined ‘social and structural barriers’ among those with co-occurring disorders (COD), that is ‘mental illness, substance abuse, and general medical conditions encounter[ed] in regard to their health care’. Social barriers were experienced by those with COD including difficulties relating to their interpersonal ‘relationships with health care providers’ and ‘negotiating an arduous the healthcare system’ (e.g. problems relating to ‘inadequate insurance coverage and medication capitations’). These challenges were compounded by wider environmental determinants, in particular ‘living in contexts that made health management difficult, such as unkempt and dangerous housing accommodations.’

Another social environment study (IX) focused on identifying household and area-level social determinants of multimorbidity and co-morbidity. Across three main social determinants of household composition, household tenure and household rurality, mixed evidence was found of these determinants being associated with either increased or decreased multimorbidity. Much of the evidence presented in this study was found to be inconclusive in relation to the role and effect of household and area-level social determinants on multimorbidity.

The third study (X) aimed to ‘determine the prevalence of multimorbidity and examine its association with various social determinants of health in South Africa.’ Variables including age, gender, education, income, employment, obesity, depression, income and smoking, were found to be associated with multimorbidity. The authors also reported that social capital (defined as ‘how people connect with others in their environment’), which has been shown in the literature to be beneficial for health, was not associated with multimorbidity or even health per se.

Another study (XI) focused on identifying ‘associations between lifestyle behavioral factors and multimorbidity resilience (MR) among older adults’. A broad range of variables were used including four social and environmental variables: number of friends, number of relatives, housing problems and urban/rural status. Results indicated all four factors ‘exhibited statistically significant, albeit weak, associations with multimorbidity’.

#### The natural environment

The natural environment was the focus of one study (XII). This explored the effects of environmental degradation (resulting from dry salinity) on mental health in relation to ‘diseases co-morbid [asthma, ischaemic heart disease, suicide/self-inflicted injury] with depression in this environmental setting’. It found that ‘the association of asthma, suicide and heart disease with salinity was most likely attributable to the co-morbidity of the conditions with depression’.

## Discussion

This rapid scoping review was conducted to identify and gain a broad overview (12) of the key research evidence on the role of wider environmental determinants in the MLTC sequelae. All but one study identified by this review considered either the built environment or social environments or both, and how these aspects of environmental determinants related to MLTC. While a relatively broad range of specific determinants were examined in the studies identified by this review, there was an analytical focus on various aspects of the quality of housing/accommodation and living conditions, external air pollution and indoor air quality, and the design and configuration of urban environments and neighbourhood characteristics (e.g. neighbourhood levels of crime, accessibility to healthcare services, availability of green spaces). These studies did find evidence of correlations between these determinants and increased or decreased risks of MLTC, particularly in relation to the quality of internal housing/living environments, exposure to airborne environmental hazards (e.g. air pollution/air quality), and a beneficial association with socially cohesive, accessible and greener neighbourhood environments. It is not clear why these aspects of environmental determinants featured more prominently in this review, although one reason might be that they are more easily quantified and measured, and data sources (including electronic health, public health and environmental health records) are more readily available and accessible to researchers. Additionally, these environmental determinants may also be more prevalent and widely reported, and as such there is wider societal awareness of these determinants, which in turn may have led to greater scrutiny and examination of possible associations with MLTC. Very little evidence was found on the role of the natural environment on established MLTC, suggesting this aspect of environmental determinants is a particularly under-researched field.

## Strengths and limitations

We carried out a rapid scoping review to summarise evidence on MLTC and environment that could inform further research in this field. We included broad inclusion criteria of MLTC and any environmental determinants to help identify a wide range of studies including grey literature. However, more specific research terms around MLTC and environmental determinants, including, for example, housing, pollution, etc; may have increased the number of papers identified. Further, we limited papers to those published in the English language which could have limited identification of all relevant papers in this area. We did not carry out quality assessment of papers as this is not a requirement of scoping reviews.

## Conclusions

This scoping review aimed to collated evidence on environmental determinants in MLTC. We identified only 12 relevant papers in this field, and most focused on the built and social environments. The evidence on the natural environment was extremely limited. Overall, the review uncovered very limited and this finding indicates that further research is needed to understand the role of environmental determinants in MLTC.

## Data Availability

All data produced in the present work are contained in the manuscript.

## Ethics approval and consent to participate

Ethical approval was not required for this scoping review and therefore not applicable.

## Availability of data and materials

Data used during the current study are available from the corresponding author on reasonable request.

## Author Contribution

All authors contributed equally. HDM is guarantor.

## Acknowledgements

None.

## Author’s information

None.

## Consent to publish

Not applicable.

## Declaration

The lead author affirms that the manuscript is an honest, accurate, and transparent account of the study being reported; that no important aspects of the study have been omitted. The opinions, results, and conclusions reported in this article are those of the authors and are independent from the funding sources.

## Appendix

**Table 1:**
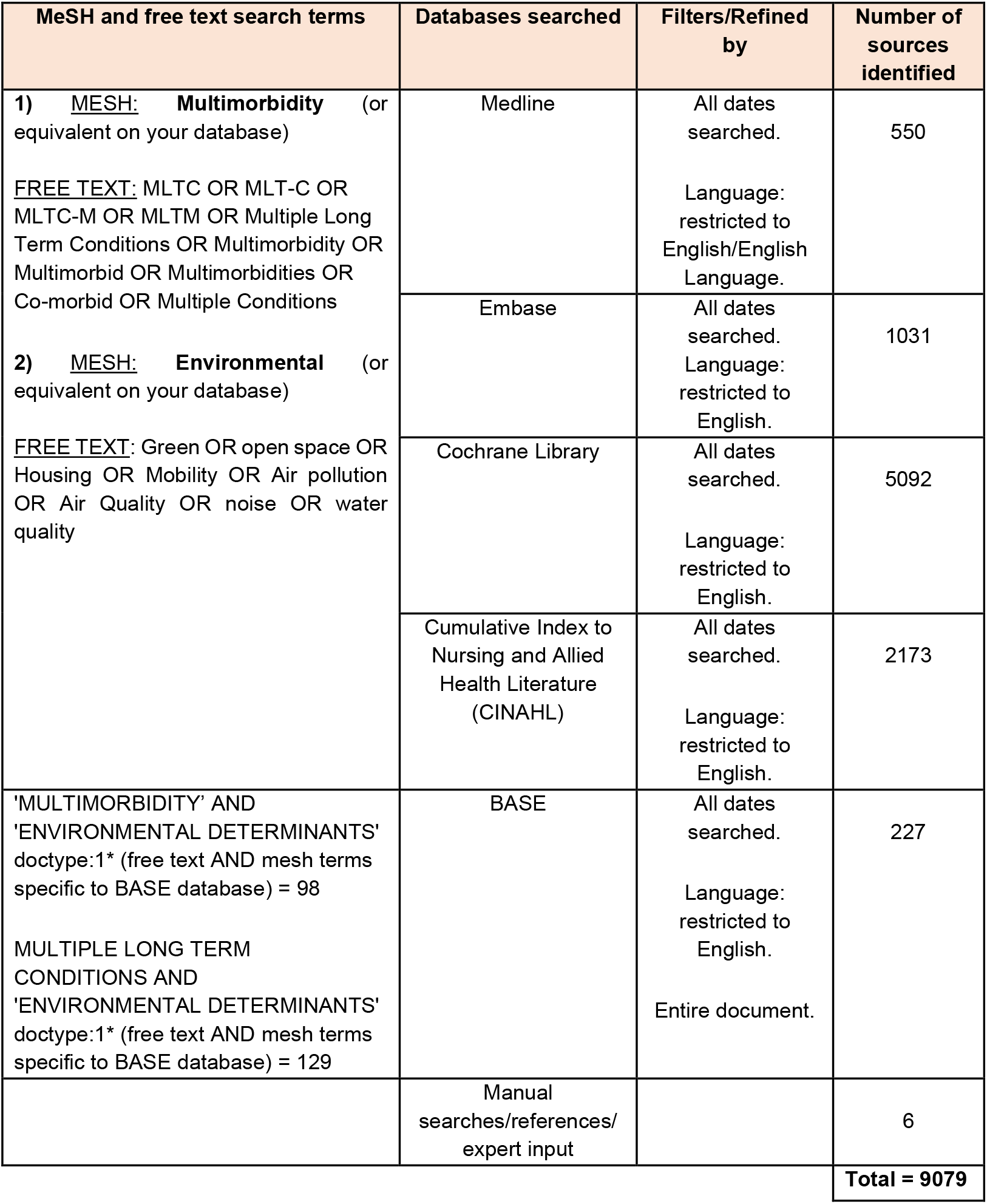
Search terms results table.

**Table 2:** Key characteristics of included studies.

| Study | Source - first author and date | Setting (e.g. primary care, secondary care) | Country (e.g. UK, USA) | Population characteristics: age, sex, ethnicity & disease group | Sample size | Study type (Qualitative, Mixed Methods or Quantitative) | Study aims | Findings (including links with MLTC) | Environmental determinants discussed |
| --- | --- | --- | --- | --- | --- | --- | --- | --- | --- |
| I | Wu, Yu-Tzu, 2020.<br>The longitudinal associations between proximity to local grocery shops and functional ability in the very old living with and without multimorbidity: Results from the Newcastle 85+ study | Community / neighbourhood setting | UK | People aged 85+<br>People with and without MLTC | A population-based cohort of 852 people | Quantitative | To identify environmental factors that support maintenance of functional ability, the aim of this study is to investigate the longitudinal associations between proximity to local grocery shops and the ability to shop for groceries in the very old and to examine the potential variation between those living with and without multimorbidity. | The very old who lived in more deprived areas were more likely to have a grocery shop within 500 m than those in less deprived areas. Proximity to local grocery shops had limited impacts on those who were relatively healthy (0-1 chronic condition), but moderated loss of the ability over time in those living with multimorbidity.<br>Proximity to local grocery shops was, however, found to support the very old living with multimorbidity and their ability to do grocery shopping. | 'Other environmental factors (safety, traffic, stairs, the quality of pavement, age-friendliness of local shops) ... might affect use of local shops.' |
| II | Adhikari, B. 2021. Community design and hypertension: Walkability and park access relationships with cardiovascular health | Community / neighbourhood setting | Canada | Two independent population cohorts of adults.<br>Hypertension and related cardiovascular disease (CVD).<br>The studies recruited participants using different age limits, the average age of participants was 45.6 years and 55.1 years for MHMC and BC Gen, respectively.<br>Ethnicity was categorized into five groups: (i) Caucasian, (ii) Aboriginal, (iii) Asian, (iv) South Asian, and (v) for MHMC and into two groups: (i) Caucasian and (ii) others for BC Gen. | One Cohort was 22,418 adults (My Health My Community) and the other cohort was 11,972 adults (BC Generations Project). | Quantitative | To examine how features of the built and natural environments relate to hypertension and the mediating role of transportation and leisure walking and body mass index in this relationship. | Findings demonstrate that living in a walkable neighbourhood and having higher park accessibility is associated with lower odds of hypertension, especially for lower income individuals.<br>We suggest an integrated population health approach that considers multimorbidity as a result of exposure to car-dependent areas and the lack of green spaces. | Neighbourhood walkability and park accessibility may help mitigate adverse health consequences of hypertension by reducing the number of individuals with body mass index. |
| III | Alkhatib, A. 2021. Preventing Multimorbidity with Lifestyle Interventions in Sub-Saharan Africa: A New Challenge for Public Health in Low and Middle-Income Countries | No specific setting | Sub-Saharan Africa | Adults aged 18+<br>Males and females.<br>Multimorbidity clusters, especially hypertension, diabetes and cardiovascular disease could provide an early effective prevention of multimorbidity in LMICs [Low and Middle Income Countries]. | Not applicable | Narrative review | This narrative review summarises key epidemiological multimorbidity determinants and highlights specific public health challenges related to multimorbidity prevention strategies, especially through lifestyle in LMICs placing emphasis on Sub-Saharan Africa, and its future regionally relevant health policies. | LMICs [Low and Middle Income Countries] are experiencing a fast-paced epidemiological transition towards multimorbidity, characterized by clusters of NCDs [Non-communicable Diseases], which require public health interventions. Multimorbidity determinants include increased age, female sex, environment, lower socio-economic status, obesity, and lifestyle behaviours, especially poor nutrition, physical inactivity. | Long term negative impact of environmental factors on health outcomes has particularly focused on air pollution associations with the development of chronic asthma, pulmonary insufficiency, CVD [cardiovascular disease], cancer, and diabetes. |
| IV | Adamkiewicz. G. 2014. Environmental Conditions in Low-Income Urban Housing: Clustering and Associations With Self-Reported Health | Low-Income Housing developments | USA | Participants from the adult population of 20 publicly and privately managed low-income housing developments across 3 cities (Cambridge, Somerville, and Chelsea) in the metropolitan area<br>of Boston, Massachusetts.<br>Residents of the developments were eligible for participation if they were aged 18 years or older and spoke English, Haitian Creole, or Spanish. | 828 | Mixed methods | We explored prevalence and clustering of key environmental conditions in low-income housing and associations with self-reported health. | Environmental problems were common; more than half of homes had 3 or more exposure-related problems. After adjustment for household-level demographics, we found clustering of problems in site for pests, combustion by products, mold, and ventilation. | Mold, combustion by-products, second-hand smoke, chemicals, pests, and inadequate ventilation. |
| V | Hernandez-Garcia, E. 2021. Research capacity strengthening in health and care delivery through housing for chronic diseases | No specific setting. | No geography specified | No specific population specified.<br>No age range specified.<br>No ethnicity specified. | Not applicable | Systematic review | The aim was to determine relevant evidence gaps to building capacity in supportive housing-health care research about chronic diseases and multimorbidity, identify ongoing strains and offer potential solutions. | Future research needs to prioritise articulating these emerging insights through systematic, translational and multisectoral approaches to strengthen health and care delivery through housing design solutions for people living with chronic morbidity. | Impact of housing design on MLTC. |
| VI | Adamkiewicz, G. 2014. Neighborhood Environment and Type 2 Diabetes Comorbidity in | Community / Neighbourhood setting | Australia | Adults over 18 years old with a "Serious Mental Illness" such as schizophrenia, bipolar disorder, | 3,816 | Quantitative | We aimed to investigate the associations of neighborhood environments with T2D comorbidity in | T2D comorbidity in SMI is a major public health issue.<br>We observed that individuals with SMI residing in areas with | Our study focused on 5 neighborhood-level variables: (1) neighborhood-level crime, (2) access to health care services, (3) neighborhood-level |
|  | Serious Mental Illness |  |  | or major depression. |  |  | individuals with SMI. | higher crime rates were more likely to report T2D comorbidity compared to individuals with SMI residing in lower crime rate areas, even after controlling for individual-level variables and neighborhood-level disadvantage.<br>Overall, the study suggests that the mechanisms of neighborhood influence on SMI-T2D are highly complex. | obesity, (4) availability of green spaces, and (5) availability of fast-food outlets. |
| VII | Ou, J. Y. 2018. Self-rated health and its association with perceived environmental hazards, the social environment, and cultural stressors in an environmental justice population | Community / People at home | USA | Adults over 18 years of age.<br>Hispanic urban population. | 354 | Mixed methods | We examined self-rated health and its association with multiple types of perceived environmental hazards in a majority-Hispanic urban population. | We identify strong associations among fair/poor self-rated health and participant-reported environmental hazards, the social environment, and cultural stressors.<br>We confirm the roles of environmental, social, and cultural stressors on self-rated health even after adjusting for chronic and mental health conditions. Our findings shed light on the complex sources of chronic stress in an<br>environmental justice community, and their impacts on the health of a majority-Latino population. | Self-rated health, perceptions of their neighborhood, including participant-reported environmental hazards (e.g., air quality, odors and noise), aspects of the social environment (e.g., feeling safe, neighborhood crime, social cohesion), and culture-related stressors (e.g., immigration status, language stress, ethnic identity). |
| VIII | Villena, A.L.D. 2010. Challenges and struggles: lived experiences of individuals with co-occurring disorders | Community / Neighbourhood setting<br>Community treatment centers and supportive housing sites | USA | Mean age 51.<br>60% with MLTC.<br>40% with one condition.<br>Psychiatric and physical diseases.<br>Participant characteristics: 11 men and 9 women; (65% African American) | 20 | Mixed methods<br>Qualitative interviews.<br>Quantitative analysis of conditions and demographics. | The purpose of this interpretive study was to understand, describe, and illustrate the social and structural barriers that individuals with COD [Co-occurring disorders] of mental illness, substance abuse, and general medical conditions encounter in regard to their health care. | Living with COD was perceived to be a “constant struggle and challenge” by informants. Their primary and pervasive complaint was that relationships with health care providers served as impediments to managing health. Informants felt misunderstood and at times voiceless.<br>In addition, inadequate insurance coverage and medication copayments created additional burdens for this population trying to manage multiple health problems simultaneously. These barriers were worsened by living in contexts that made health management difficult, such as unkempt and dangerous housing accommodations. | Navigating healthcare system.<br>Trust with providers (doctors).<br>Quality of accommodation/housing including stability.<br>Support from family and friends. |
| IX | Ingram, E. 2020. Household and area-level social determinants of multimorbidity: a systematic review | Community / neighbourhood | North America, Europe and Australasia | Participants from the general population and assessed for the presence of multiple chronic conditions (multimorbidity).<br>Participants solely young people (age <18 years) excluded. Age range mainly those aged 50 and over.<br>Thirty-six of 41 studies included a mix of chronic physical and mental health conditions, while four included physical conditions only. | Not applicable | Systematic review | This review aimed to systematically identify, critically appraise and synthesise the existing literature on associations between household and area-level Social Determinants of Health and multimorbidity prevalence or incidence.<br>In general populations of high-income countries (HICs). We also aimed to investigate how associations<br>Differ with age, gender and ethnicity. Better understanding of SD of multimorbidity could inform equitable prevention and intervention strategies. | Those living in the most deprived areas had the highest prevalence or incidence of multimorbidity.<br>Associations between deprivation and multimorbidity differed by age and multimorbidity type.<br>Findings from the few studies investigating household tenure, household composition and area-level rurality were mixed and contradictory; homeownership and rurality were associated with increased and decreased multimorbidity, while living alone was found to be associated with a higher risk of multimorbidity and not associated. | Household tenure, household composition and area-level rurality.<br>Trust in neighbours (n=1): One fairly low-quality study found that participants who somewhat distrusted their neighbours had increased risk of developing multimorbidity within 11 years compared with those who strongly disagreed with the statement 'One cannot trust each other here' (RR 1.13, 95% CI 1.03 to 1.23). |
| X | Olufunke A. 2013. The social determinants of | Community / People at home | South Africa | Adults over 18 years of age. | 11,638 | Quantitative | The aim of this study was to determine the | Social capital (defined as 'how people connect with others in their | Social capital |
|  | multimorbidity in South Africa |  |  | Males and Females.<br>(Surveys were given to all above 15 but those below 18 were removed).<br>A diverse range of ethnic groups in a South African context. |  |  | prevalence of multimorbidity and examine its association with various social determinants of health in South Africa. | environment'), was not associated with multimorbidity or even health per se. |  |
| **XI** | Health behaviors and multimorbidity resilience among older adults using the Canadian Study on Aging | Community / People at home | Canada | Canadian adults aged 65 or older who reported two or more of 27 chronic conditions.<br>Conditions included:<br>Alzheimer's disease, back problems, bowel incontinence, cancer, cataracts, diabetes, epilepsy, glaucoma, heart attack, heart<br>disease, high blood pressure, irritable bowel syndrome, kidney disease, Parkinson's disease, peripheral vascular disease, lung disease, macular degeneration, multiple sclerosis, osteoarthritis, osteoporosis, migraine headaches, rheumatoid arthritis, stroke, thyroid problem, transient ischemic attack, ulcer, and urinary incontinence. | 6,771 | Quantitative | This study examines associations between lifestyle behavioral factors and multimorbidity resilience (MR) among older adults. | All four of the social/environmental variables exhibited statistically significant, albeit weak, associations with multimorbidity. | Social and environmental variables: Number of friends, number of relatives, housing problems and rural/urban status. |
| **XII** | Speldewinde, P.C. 2011; The hidden health burden of environmental degradation: disease comorbidities and dryland salinity | Community (rural) | Australia | The case populations with the three diseases (asthma, suicide and ischaemic heart disease) studied between 1996 and 2001, were extracted | Total population of the study area averaged 408,111. | Quantitative | Given the emerging evidence of the effects of environmental degradation on mental health, it was hypothesised that typically comorbid diseases | Using georeferenced health record data, Bayesian spatial methods were used to determine the relationship between dryland salinity and a range of human health outcomes. Initial | The effects on health of environmental degradation related to dry salinity. |
|  |  |  |  | from Western Australia's Data Linkage Unit database.<br>Indigenous people.<br>Adults 18+. |  |  | may also be influenced by environmental degradation. In Western Australia, a recent study (Speldewinde et al., 2009) found an association between dryland salinity and depression. This rural population was re-examined for a range of other physical and psychological conditions—particularly asthma, ischaemic heart disease and suicide—to identify whether any corresponding increased risk was associated with the presence of a depressive illness. | modelling found an increased relative risk for asthma, suicide and ischaemic heart disease in relation to dryland salinity (adjusted for Indigenous and socio-economic status). However, in this follow-up study, a further evaluation of the role of co-morbidities in this population revealed that: (i) the presence of depression was consistently linked to residence in areas with high salinity and (ii) the association of asthma, suicide and heart disease with salinity was most likely attributable to the co-morbidity of the conditions with depression. |  |
|  |  |  |  |  |  |  |  | **Total included** | 12 |

